# An Adaptive Technical and Behavioral Approach for Biomarker Acquisition in Autism with Intellectual Disability

**DOI:** 10.64898/2026.10.01.26364439

**Authors:** Cassie Stevens, Sara Eberle, Casey Carrow, Sherab Tsheringla, Christine Cukar-Capizzi, Julie Wolf, Adam Naples, James C. McPartland

**Affiliations:** Yale Child Study Center, New Haven, Connecticut; Schiefelbusch Institute for Life Span Studies, University of Kansas, Lawrence, Kansas; Yale Center for Brain and Mind Health, New Haven, Connecticut

**Author notes:** Correspondence concerning this article should be directed to James C. McPartland, Ph.D Yale Child Study Center New Haven, CT.

**Keywords:** autism spectrum disorder, intellectual disability, profound autism, severe autism, EEG, eye tracking, biomarkers

## Abstract

Autistic individuals with intellectual disability (ASD+ID) and profound autism are rarely studied in neuroscience research due to barriers including communication challenges, sensory sensitivities, anxiety, and elevated behavioral support needs. Consequently, biomarker development in autism fails to include the portion of the autistic population with highest support needs and greatest potential to benefit from treatments derived from imminent advances in personalized medicine. Here we present a multimodal, participant-centered protocol for simultaneous electroencephalography (EEG) and eye-tracking (ET) acquisition to advance clinical trial readiness research in this underserved population. The protocol combines individualized behavioral supports and an adaptive EEG-ET acquisition platform. Pre-visit caregiver interviews informed participant-specific research plans, including customized visual supports, reinforcement strategies, and EEG net desensitization procedures. A gaze-contingent stimulus presentation system used participant-preferred videos to support attention to experimental stimuli and behavior consistent with data acquisition (e.g., refraining from movement) while dynamically adapting stimulus delivery based on visual engagement with the assay and real-time assessment of data quality. By quantifying attended trials in real time and prioritizing unattended stimulus classes, the paradigm maximizes acquisition efficiency to minimize participant burden. This approach was utilized to deploy candidate biomarkers with demonstrated replicability in the Autism Biomarkers Consortium for Clinical Trials (ABC-CT): the N170 event-related potential to faces, resting-state EEG, and visual attention to human faces (measured with ET). Feasibility was evaluated in 76 children with ASD+ID (M age = 8.76 years; IQ = 32.4) and a small comparison sample of 8 children with intellectual disability without autism (M age = 8.91 years; IQ = 45.9). Usable EEG data (>7 minutes) were acquired in 61% (ASD+ID) and 88% (ID) of participants, respectively. Sufficient ET data were obtained from 67% (ASD+ID) and 63% (ID) of participants, respectively. Findings demonstrate the feasibility of inclusive biomarker acquisition and provide a framework for expanding participation of individuals with ASD+ID and profound autism in neuroscience research. Ongoing research seeks to evaluate this approach in larger samples and to demonstrate feasibility in multisite research studies.

## Introduction

The majority of autistic individuals have at least one co-occurring medical, psychiatric, or neurodevelopmental diagnosis (e.g., ADHD) (Hossain et al., 2020; Micai et al., 2023). Intellectual disability (ID), a neurodevelopmental condition characterized by significant limitations in intellectual and adaptive functioning, is one of the most common co-occurring conditions yet is critically understudied in autism research. Approximately 38% (Maenner et al., 2023) of autistic individuals have co-occurring ID, and 27% are estimated to have profound autism (Hughes et al., 2023). However, only 6% of published autism studies include participants with ID (Russell et al., 2019), and recent meta-analyses of electroencephalographic (EEG; Kang et al., 2018) and eye-tracking (ET; Frazier et al., 2017) indices of social-communication in school-aged children indicated that only 2 of 145 studies focused on autistic samples with ID (ASD+ID). As such, scientific knowledge of these subgroups is sparse, and the subgroup of autistic individuals with the highest support needs are being excluded from potentially care-advancing research. Advancing etiological understanding and developing precision interventions for those with ASD+ID will require both an intentional research focus and flexible data collection methods accommodating of this group’s needs.

Autistic individuals with ID face numerous barriers to inclusion in research at all stages of study participation, from recruitment through successful completion of study procedures. Challenges to participation in neuroscience research include: 1) limited functional communication and difficulty following complex verbal instructions, 2) difficulty sitting still and attending to standardized stimuli for sustained periods of time, 3) anxiety with unfamiliar people, settings, and equipment, and 4) challenging behaviors such as aggression, self-injury, and property destruction (for general review, see Maes et al., 2021). To address these barriers, research protocols must include tailored strategies to support comfort and engagement without compromising scientific rigor.

EEG and ET methods are well-suited for biobehavioral measurement in ASD+ID, as these methods require minimal verbal or motor responses, and are quieter and more robust to participant motion relative to other neuroimaging methods (Jack & Pelphrey, 2017). ET, and particularly passive viewing ET paradigms, place minimal demands on participants and have been successfully used with autistic individuals with ID and minimally verbal autistic children (e.g., Plesa Skwerer et al., 2016; Tager-Flusberg et al., 2017). Furthermore, EEG and ET methodologies offer significant potential public health benefits, as they are more scalable and economical than most neurobehavioral acquisition techniques. Prior research has demonstrated the effectiveness of social stories, video modeling, stakeholder-informed procedures, and EEG net desensitization in obtainment of high-fidelity EEG and ET data in ASD+ID (DeMore et al., 2009; DiStefano et al., 2019; Guy et al., 2021; Kuschner et al., 2021; Slifer et al., 2008). However, these approaches have generally relied on static experimental paradigms with limited individualization of behavioral supports during data collection. There is a critical need for innovative biomarker acquisition methods to address these challenges and increase the validity and reliability of signals in individuals with ASD+ID.

Here, we present a multi-modal acquisition protocol that integrates participant-specific preparation, clinician-guided behavioral support, and a novel, participant-responsive hardware and software apparatus to simultaneously collect EEG and ET data in ASD+ID and an ID only comparison group. Prior EEG studies in this population have often relied on resting EEG or auditory assays that do not require a visual focus to be maintained. Technical adaptations in this protocol expand upon prior work to enable acquisition of EEG to visual stimuli, allowing for the application of some of the ET and EEG biomarkers most widely utilized in studies of ASD without ID. Behavioral support strategies were informed by stakeholders, developed *a priori* with each participating family, and refined with day-of visit experiences. By combining clinical expertise, principles of applied behavioral science, and responsive experimental software, this approach aims to reduce participant burden while maximizing the efficiency and quality of biomarker acquisition in a population that has historically been excluded from neuroscience research. We detail specific technical and behavioral support components and provide feasibility data on their successful implementation in ASD+ID, providing an overarching framework for future neuroscientific research in this group.

## Methods

The present protocol for collecting simultaneous EEG-ET data was developed to complement the protocols and assays from the Autism Biomarkers Consortium for Clinical Trials (ABC-CT; McPartland et al., 2020) and with a programmatic commitment to include individuals with ASD+ID in biomarker research. Our adaptations addressed the specific needs of individuals with ASD+ID across the data collection workflow, from pre-visit preparation through data collection (Figure 1). Procedures were refined in collaboration with the Yale Autism Program Community Council, composed of autistic adults and caregivers of autistic people to ensure practices are aligned with lived experiences of community members.

**Figure 1.**
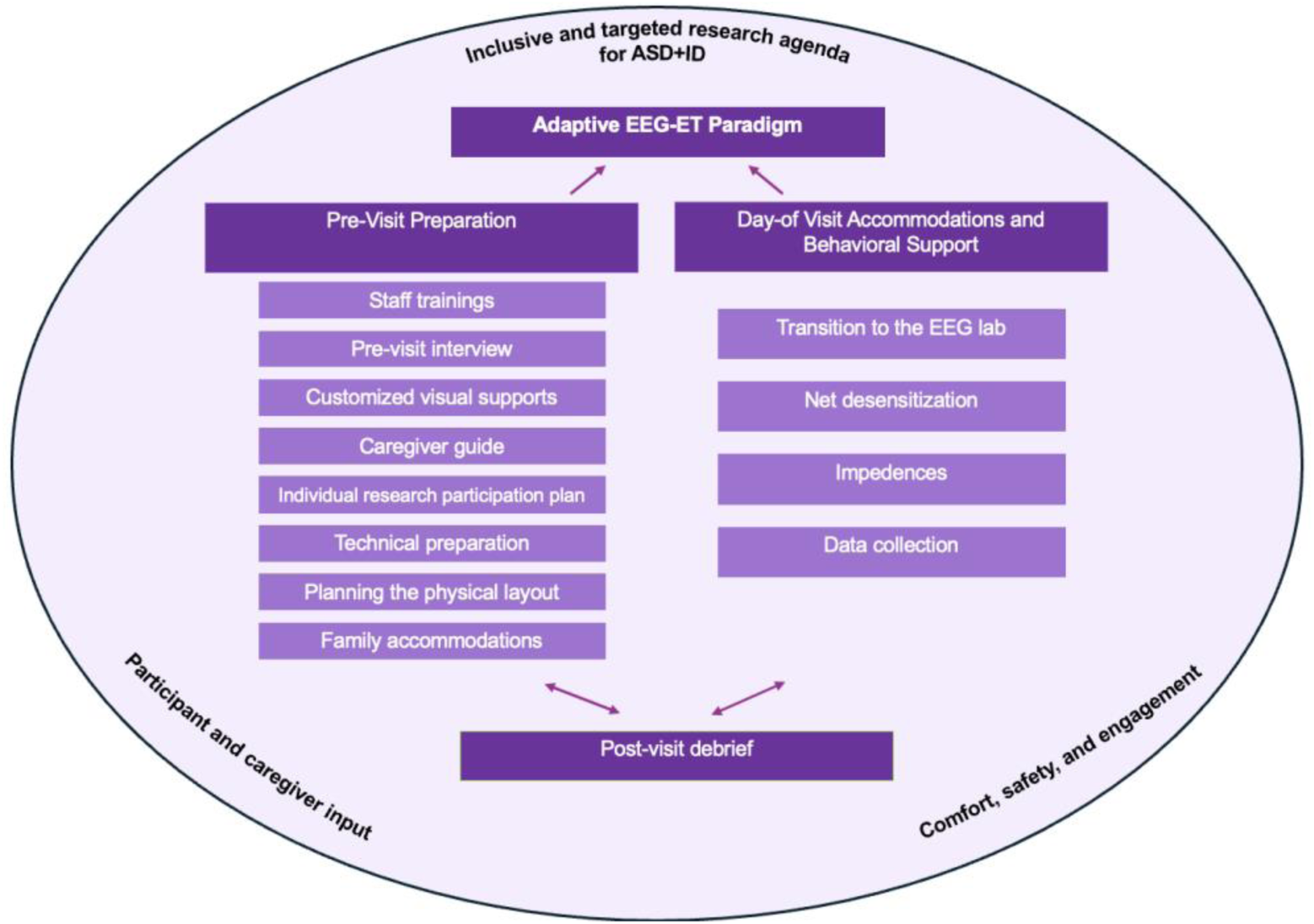
Summary of framework for inclusive EEG-ET research protocol in an ASD+ID sample.

The protocol was designed to be implemented by two research staff and a clinical expert (i.e., a clinical psychologist and/or BCBA). The two research staff included (1) an experiment operator, who operated the experimental presentation system and ET-EEG data collection and (2) a behavioral support staff member, who facilitated participant engagement and completion of study procedures. When staffing permitted, a second behavioral support staff member could be included based on the anticipated or observed behavioral support needs of the participant. The clinical expert provided behavioral training to research staff and clinical oversight of visit preparation and participant support throughout visits.

First, we developed pre-visit preparation approaches for data collection for staff, participants, and families. Second, we adjusted the physical layout of the experimental space and participant-facing materials (e.g., social stories) to anticipate participant and family needs. Third, we adjusted protocols for working with children during data collection, particularly for activities such as EEG net application and ET calibration. Finally, we modified our experimental delivery system to be more flexible to facilitate biomarker data collection. Below, we describe these modifications.

Participants were clinically characterized at a separate visit by one of the team’s clinical experts. IQ was measured using the Differential Ability Scales Second Edition (DAS-II) School-Aged and Early Years forms (Elliott, 2007) and the Mullen Scales of Early Learning (Mullen, 1995) when appropriate. The best available full-scale IQ was used. Participants were also administered the Autism Diagnostic Observation Schedule, Second Edition (ADOS-2; Lord et al., 2012) for diagnostic confirmation. The study was performed under a protocol approved by the Yale University Human Investigation Committee. Written informed consent was obtained from all caregivers after a complete explanation of study procedures, including clarification that discontinuation of participation would not yield negative consequences. Families were offered a written report and feedback with an expert clinician as part of participation.

### Pre-Visit Preparation

#### Staff trainings

Expert clinicians trained all research staff on behavioral principles and strategies to support implementation and confidence in running study visits. Training covered proactive visit preparation, application of behavioral techniques, and “on-the-fly” decision-making for best visit outcomes. Visit preparation training included environmental and situational modifications made before behavior occurs to increase the likelihood of desired behavior (i.e., antecedent controls). Behavioral support staff learned strategies for differentially reinforcing desired behaviors and approximations (i.e., partial or successive steps that increasingly resemble desired behaviors). Staff were also trained to respond calmly and minimally to challenging behavior while prioritizing participant safety and rapidly reinforcing successful task engagement. Staff first practiced behavior support strategies in role-plays with an expert clinician and then with one another. When interacting with participants, staff used simple, concrete language.

### Pre-visit interview

Research staff conducted a virtual pre-visit interview with caregivers to characterize each child’s developmental abilities, communication style, sensory sensitivities, history of challenging behaviors, interests, and reinforcers. Caregivers often offered to bring items from home, and caregivers and staff collaboratively decided which reinforcers, break time activities, snacks, or comfort items might be supportive on visit day. When available, an inventory of any functional behavioral assessments, established behavior plans, and currently effective behavior management strategies were collected prior to the research visit.

The team also collected suggestions of preferred videos and music their child found motivating to integrate into the EEG-ET stimulus presentation. Care was taken to exclude media that may increase motion (e.g., dancing, repetitive physical behaviors). Caregivers were interviewed regarding effective strategies to sustain the child’s interest and attention. Caregivers were provided an overview of visit activities and asked to anticipate any challenges that may result from the visit environment or study activities. The caregiver interview guided staff in creating tailored and developmentally informed research participation plans prior to visits (see Appendix A for full interview).

### Customized visual supports

Successfully implemented visual supports for our study participants included personalized social stories, first/then boards, token boards, checklists, and visual schedules. First/then boards presented a preferred activity or item that would follow completion of a less-preferred task (e.g., “First put on hat, then have snack”), while token boards allowed children to earn stars or tokens toward a preferred prize or reinforcer for completing designated tasks. Visual supports were customized based on child preferences and report of previously effective strategies. Caregivers were sent personalized social stories and other visual supports for the planned research visit in advance, and printed copies were provided upon arrival to the research visit. Information about the tailoring of these supports, including customizable templates, can be found in Appendix B.

### Caregiver guide

After the pre-visit interview, caregivers received a guide to set expectations for the visit, support participant engagement, and maximize fidelity to the EEG/ET protocol. The guide described the purpose and setup of an EEG, caregiver guidance for helping child adjust to wearing the EEG net (i.e., net desensitization), and guidelines for providing behavioral assistance during the EEG session when applicable (Appendix C). A laminated, abbreviated version of the guide was also available in the lab space as a caregiver visual support during the visit (Appendix D).

### Individualized research participation plan

Behavior support staff met with one of the clinical psychologists and/or BCBAs on the research team to develop an individualized participation plan based on the pre-visit caregiver interview responses and any additional supporting information (e.g., IEP, behavior plan, recent psychological testing). For each participant, staff and caregivers formulated a plan to help the individual be desensitized to wearing the EEG net. Desensitization plans specified where desensitization would begin (e.g., at home, in the waiting room, in the EEG room) and which early strategies to implement (e.g., wearing hats or getting their head wet at home, touching and/or wearing the model net). Netting plans were adapted based on participant state and response on the day of the visit (see *Net Desensitization* below). To improve tolerability, EEG nets were warmed in a water bath system prior to all visits and maintained at a consistently warm temperature before net application.

### Technical preparation

Following the pre-visit caregiver interview, participants’ preferred video clips were embedded directly within the EEG-ET paradigm. All clips were resized to so that they would fill the center of the screen to engage participant looking. Prior to participant visits, staff previewed individualized paradigms in their entirety to ensure successful integration of the videos into the experiment.

### Planning the physical layout

The EEG data collection room was arranged to reduce distractions and direct attention to the experimental stimulus display. In advance of visits, doors to other rooms in the suite were closed and distractions minimized. Planning of the physical layout was also completed (Figure 2) to determine appropriate placement of personnel, and a caregiver if needed, in the EEG study room.

**Figure 2.**
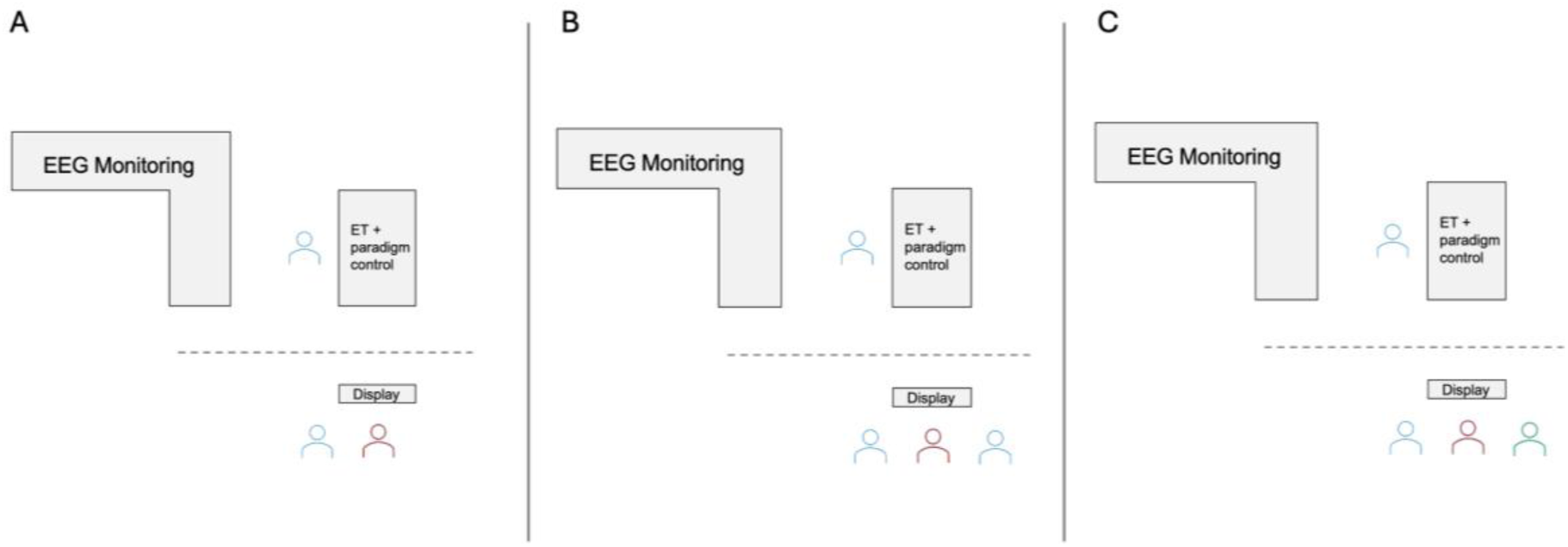
Example experimental room set-up for spatial planning. Participant location is shown in red while staff (blue) are distributed in the space for behavioral support and experiment operation. The location of the EEG monitor and ET/stimulus control stations are fixed, with curtains (dashed gray line) hanging to visually isolate the stimulus presentation monitor from the equipment and limit distractions. In all arrangements, there is one experiment operator (experimental paradigm control and ET data collection), and one (A) or two (B) behavioral support staff sitting alongside the participant. In some instances (C), a caregiver (green) provided behavior support in addition to a staff member.

### Family accommodations

Families of individuals with ASD+ID and profound autism face unique stressors and high levels of unmet need (e.g., Ferguson et al., 2025). Accordingly, the research team offered additional supports for families including evening and weekend appointments; parking passes for a connecting parking garage; coordination of snacks and lunches for families on the day of the visit; a private break room; and sibling childcare when needed. Study coordinators collaborated with caregivers and took steps to meet individual families’ additional needs whenever possible.

### Day of visit accommodations and behavioral supports

Research participants and caregivers arrived to the visit with any previously selected reinforcers or preferred items. Although tailored behavioral plans guided each visit, accommodations were refined in real time based on the participant’s day-of needs and state (e.g., fatigue, irritability).

### Arrival to research suite and consenting

Families were first greeted by behavioral support staff. The child was encouraged to freely explore the waiting room toys and activities to acclimate to the space without demands. During this period, behavioral support staff informally assessed affect, sensory sensitivities, and comfort with physical interaction. A second staff member joined the group to obtain informed consent and discuss any day-of factors that might affect EEG tolerance. Behavioral support staff obtained informed assent and measured head circumference for EEG net sizing while continuing to assess the participant’s readiness.

### Transitioning to the EEG lab

Following consent, behavioral support staff introduced and reviewed custom supports and read the social story to reiterate the sequence of visit events. Staff described the EEG session in a positive tone and emphasized that the participant would wear “a hat” with “sponges” while watching a preferred video and “other pictures.” Participants were reminded of reinforcers or rewards involved in completing steps of the EEG process. Participant responses during this period informed final adjustments to the net desensitization plan, which was initiated either in the waiting room or the EEG room.

Behavioral support staff provided transitional objects such as toys, prizes, and reinforcers that the participant was able to bring to the EEG room provided they would not interfere with the EEG equipment (e.g., sticky slime) or be distressing to part with. Additionally, every attempt was made to leave personal devices such as phones and tablets in the waiting room, so as not to have competing screens and discourage interest in the preferred videos displayed by our system. Bathroom breaks were encouraged prior to initiating the netting process to avoid disruptions during the EEG session.

Each participant’s preferred video and/or music clips played as they entered the EEG room to capture attention and make the space inviting. Curtains hung on either side of the stimulus presentation monitor to minimize the visual presence of machines and other distractors. The child was given a few minutes to watch their preferred video clips and acclimate to the space without demands. Participants were seated in the most supportive arrangement for their age, needs, and experimental requirements (e.g., booster seat, or caregiver lap) while minimizing opportunities for excessive movement.

### Net desensitization

Net desensitization was a flexible, participant-paced, graded exposure process that was dynamically adjusted throughout the visit (Table 1). Depending on participant needs, gradual changes could be made to objects touched (e.g., towels, tape measure, mock net, real net), quality of the object (i.e., dry or damp), duration of touching, or proximity of object to head to progress toward full net placement. Preferred toys or activities were incorporated when appropriate to increase engagement (e.g., having a hand puppet “help” place the net), and both praise and reinforcement were delivered for engagement in the process. More enthusiastic praise was given for progressing to successive stages of desensitization and for closer approximations to net wearing.

**Table 1.** Examples of individualized netting plans used for three different participants with ASD+ID, building on the general approach described in Figure 3.

|  | Participant A | Participant B | Participant C |
| --- | --- | --- | --- |
| **Net Desensitization** | Review social story before visit and in waiting room. | Review first/then boards before visit and in waiting room. | Review social story before visit. Token board is available in the waiting room. |
|  | Staff brings plastic model EEG net to waiting room for participant to look at and gently explore with hands. | Staff encourages play exploration with the plastic model EEG net and netting materials, including tape measure, gloves, and water bucket in the waiting room. | Staff brings dry net to waiting room. Participant explores dry net. Staff touches dry net briefly to participant's arm, then head, providing verbal praise after each attempt. |
|  | Participant holds a preferred toy brought to lab while staff applies plastic model EEG net to toy. | Staff refer to the EEG net as a "fun" or "silly" hat the participant gets to wear and provide positive reinforcement when the participant exhibits spontaneous interest in the net. | Staff applies dry net to participant's parent, clapping and celebrating with the parent and participant once applied. |
| **Net Application** | Participant holds netted toy in lap while staff nets participant. | Staff reviews first/then board with participant, first netting the participant, then providing a prize. | Staff “practices” netting the participant by touching a wet net to the participant’s head repeatedly. |
|  | Participant uses pipette to wet the electrodes on the toy’s net while staff wet the electrodes on participant’s net. | Participant needs breaks during the process of wetting electrodes. Preferred videos, coloring, and prizes are available. | Once participant is relaxed, staff stretch net over participant’s head. Participant needs a break from net. Staff removes net and tries again. |
| **Net wearing** | Staff supply preferred videos, verbal praise, and hand squeezing to support participant. | Staff provide verbal encouragement, preferred snacks, and offer coloring breaks during the paradigm. | Staff move quickly through experimental process with minimal interruption, offering slightly fewer breaks in order to maintain participant attention. |

In the waiting room, staff first encouraged the participant to touch a dry, model plastic net with their hands, followed by the model net touching the participant’s head. After transitioning to the EEG room, staff encouraged the participant to gently touch the wet net with their hands, followed by the wet net touching their head, and finally, at the participant’s pace, the fitting of the EEG net over the participant’s head. To ensure efficient and precise placement of the EEG net with minimal post-placement adjustments, research staff utilized a two-person netting technique. While one staff member stood in front of the participant to guide the EEG net over the head, a second staff member stood behind the participant, supporting the back of the net and aligning the ear holes for proper fit. When net grabbing did occur, staff supported participants by placing their hand on the participant’s hand and applying gentle pressure towards the participant’s head before removing the participant’s grasp on the net to minimize damage to the net. Figure 3 depicts strategies used across stages of netting to support successful net wearing.

**Figure 3.**
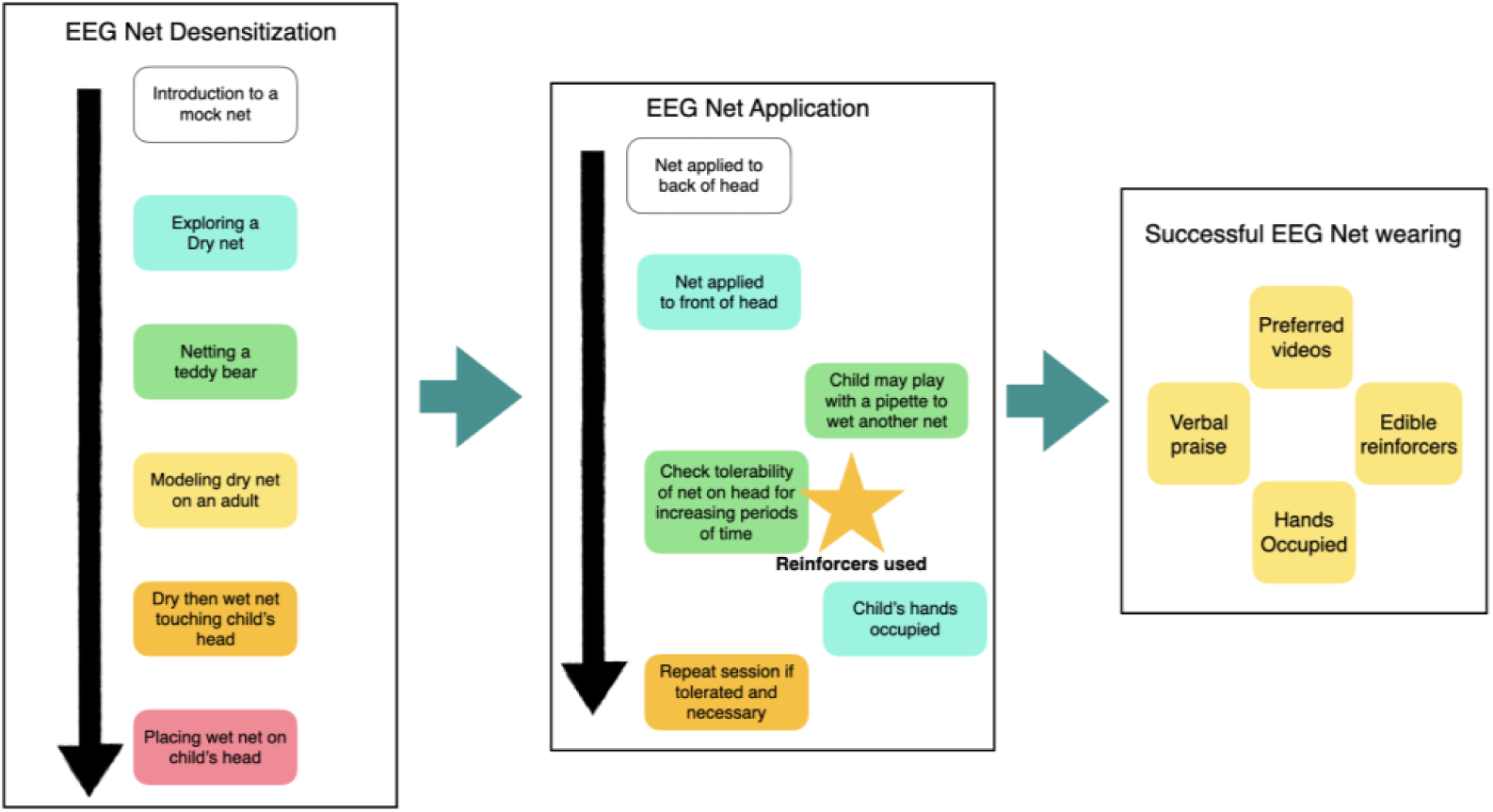
Example approaches to successful EEG net in participants with ASD+ID.

Importantly, for participants whose parents indicated that too much advanced discussion elicited anticipatory anxiety, staff were cautious not to prolong the pre-EEG period and net desensitization procedure. Staff minimized conversation or repeated exposure to the EEG net before application, instead relying on brief explanations and visual supports. This approach is consistent with prior work suggesting that limiting anticipatory exposure may reduce anxiety and improve engagement for some children (Guy et al., 2021; Webb et al., 2015).

### Impedances

EEG data were collected using 128 electrode EGI HydroCel Geodesic Sensor Nets. Adjusting impedances following net application requires parting the hair to ensure electrode sponges reach the scalp and re-hydrating electrode sponges and similar de-sensitization strategies were applied. Staff wet or re-positioned electrodes in short bursts and provided frequent breaks. Since re-hydrating all 128 electrodes may be a time-consuming and sensory-demanding process, staff sought to balance signal optimization with participant comfort.

Reference, ground, and electrodes essential to planned analyses were prioritized (e.g., those comprising the montage from which key derived variables would be extracted). If participants showed signs of behavioral or emotional dysregulation, data collection would commence even if impedances were higher in non-essential scalp regions. Throughout this process, preferred videos and reinforcement strategies (e.g., verbal praise, snacks, or stationary activities, including parallel pipette and water play with a demonstration net) were used to maximize engagement while staff worked efficiently to obtain the best possible signal with minimal participant burden. In this way, staff struck a real time balance between optimization of recording conditions and participant experience.

### Adaptive EEG-ET paradigm

Data acquisition leveraged co-registered ET and EEG in a gaze-contingent paradigm designed to facilitate high-quality data collection. The paradigm used reactive programming to continuously monitor participant attention and motion and adapt stimulus delivery in real time without the need for verbal instruction.

Preferred videos played in continuous clips during inter-trial intervals and paused automatically whenever gaze was not detected on-screen or a participant’s movement exceeded a set threshold. Video clips resumed playing once gaze returned to the screen and/or movement reduced. In this way, preferred video access served as reinforcement that was contingent upon the participant’s stillness and attention to the screen, effectively shaping behavior supportive of data acquisition without experimenter intervention. The experiment operator could toggle between several of the child’s preferred videos to avoid boredom and make the session maximally engaging.

As the paradigm proceeded, the experiment operator received real-time feedback on the number of valid trials per stimulus condition via an experimenter dashboard with a trial count for each stimulus condition. This enabled the experiment operator to selectively present stimuli from categories for which insufficient data had been gathered. The experiment operator could manually cue “video breaks” with a keystroke when needed to support engagement. This adaptive trial selection reduced unnecessary stimulus repetition, maximized efficiency of acquiring usable EEG and ET data, and minimized participant burden by ending conditions once sufficient quality data had been obtained.

### Data collection

Experiments began with participants seated 65 cm from a 22-inch monitor. Binocular eye movements were recorded at 500 Hz with an SR Research EyeLink 1000+ eye tracker. EEG data were recorded at 1000 Hz using EGI NetAmps 400 high-impedance amplifier and 128 electrode EGI HydroCel Geodesic Sensor Nets. Stimulus presentation was controlled through PsychToolbox stimulus presentation software. Participant video was captured for offline review of movement and affective state. Participant movement was monitored by the ET system head tracker in real-time (500 Hz) using a nickel-sized target sticker placed on the participant’s net or forehead.

The aims of the study were to examine the N170 event-related potential (ERP) to faces and visual social attention, which have been well documented to differ in ASD (Chita-Tegmark, 2016; Daluwatte et al., 2013; Kang et al., 2018) and are biomarkers included in large-scale biomarker studies in ASD (Loth et al., 2017; McPartland et al., 2020). The combined paradigm featured trials consisting of faces (3 distinct faces) and houses (3 distinct houses), as well as static social scenes (8 distinct scenes). The experiment kept track of experimental trials where participant’s attention was off-screen and would re-present trials with a target of 75 face/house trials each and 2 presentations of each static social scene. Task-free ‘resting-EEG’ was recorded during preferred video breaks.

The experiment began with a participant watching a preferred video (A). When the participant was sufficiently still and visually fixating on the screen (as measured by ET), a face N170 or ET trial was presented (B). Inset image (C) shows the individual trial structure (650ms fixation crosshair, 500ms stimulus, 600ms inter-trial interval). As such, the adaptive paradigm differentially reinforces calm sitting and attending to the screen.

Our adaptive experimental paradigm effectively supported quality data acquisition in the context of participant motion and inattention. The efficacy of this approach is illustrated in Figure 4 through representative time courses of two separate participants with differing attentional patterns. Periods of prolonged inattention such as noted by period (a) indicate time when the paradigm was primarily paused. Sections such as period (b) indicate time when trials were intermittently presented between periods of inattention. Time courses resembling period (c) show the consecutive presentation of stimuli when the participant attended for a prolonged stretch of time.

**Figure 4.**
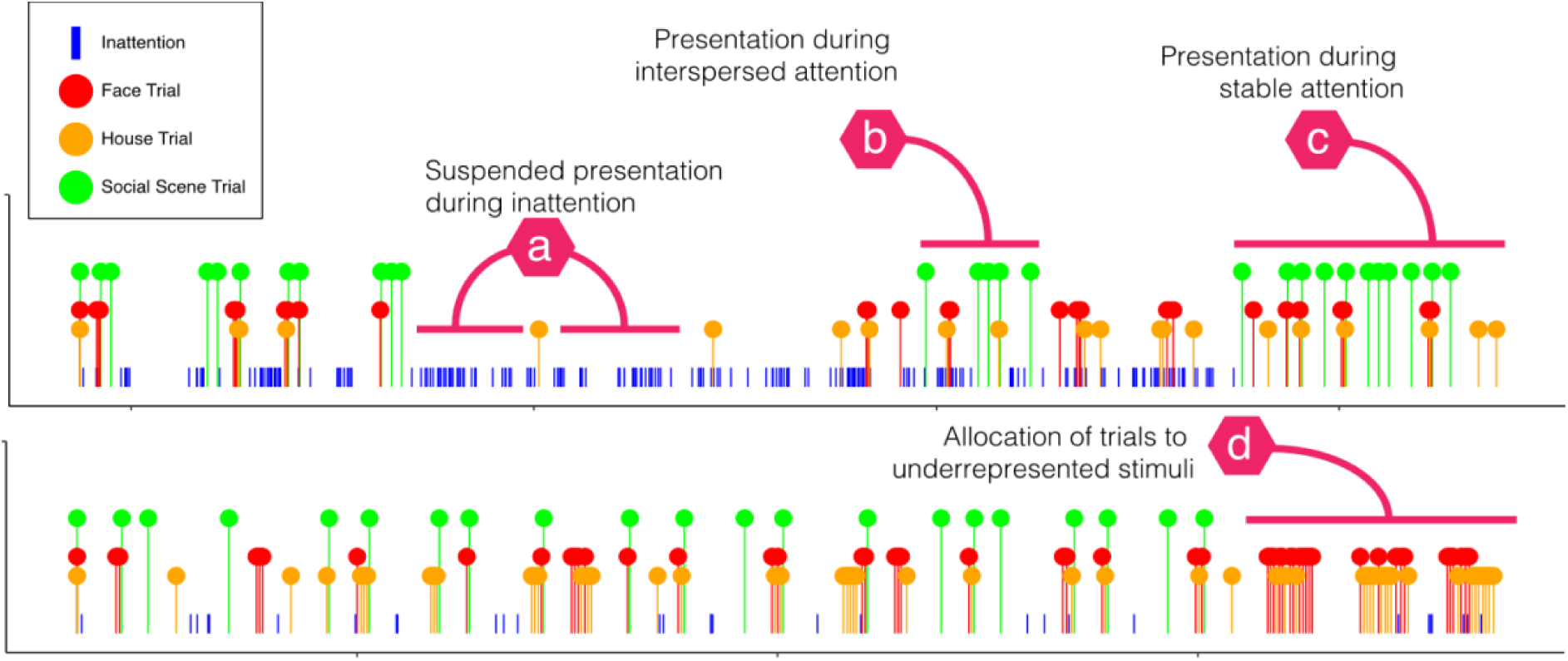
Schematic of adaptive stimulus presentation in the context of variable attention. Top panel highlights an inattentive participant, and bottom highlights an attentive participant. Colored circles represent the timing of trials by stimulus type, while blue vertical lines indicate periods of participant inattention (i.e., significant movement or looking off screen).

For the second participant, period (d) represents successive trials of face and house stimuli selectively after a sufficient number of quality social scene trials have been collected. This represents successful adaptation of the paradigm to the participant’s attentional and movement patterns and therefore maximized efficiency of data collection.

### Participant comfort and safety considerations

Participant comfort and safety were continually evaluated throughout data collection based on participant, caregiver, staff, and clinician feedback. Given the novel sensory experience of wearing an EEG net, staff prioritized a sensory-friendly environment. Some participants were provided with additional regulatory sensory input, such as having their hands held at request, or their hands and arms gently squeezed. Weighted blankets, weighted vests, and lap pads were available to provide additional sensory input and reduce movement.

Behavioral support staff reviewed caregiver-reported behaviors, common antecedents of behaviors, and effective supports before each visit. They used individualized antecedent controls, including choices, preferred activities, and visual schedules and transition plans. Staff also used safety-conscious attire, securing hair away from the face, removing dangling jewelry and ID badges to limit opportunities for grabbing or entanglement. Crisis management procedures were developed for aggressive or self-injurious behaviors, and staff were trained in de-escalation, behavioral redirection, and safe blocking. A supervising clinical psychologist and/or BCBA was available in the laboratory suite for additional support. In the event of distress or challenging behavior, staff reduced demands, provided breaks and opportunities for self-regulation, and used individualized strategies developed with caregiver and clinician input. In this non-clinical research setting, staff deferred to caregiver judgment regarding behavioral management.

While all efforts were made to keep the EEG net placed on the child’s head, it was removed if it caused continued distress to the child. When possible, staff attempted to continue ET data collection without EEG. Discontinuation of the study session was a collaborative decision between staff, clinicians, and caregivers.

### Post-Visit Debrief

Following each research visit, research staff and the supervising clinician conducted debrief discussions to evaluate which factors of the visit (e.g., behavioral support from staff, participant behavior) impacted the quality of the session. These observations were systematically documented in a post-visit “Debrief Run Log” to provide objective and quantifiable data about the child’s behavior (e.g., activity level checkboxes, instances of leaving the room, and number of netting attempts; see Appendix E) for analysis. The run log also included descriptive information about the session, such as rating of the effectiveness of staff responses to child’s behaviors and a qualitative notes section for additional context. In this way, the study continued to be participant-led, with each participant’s strategies iteratively informing the evolving toolbox of behavior management strategies.

The study team also debriefed with participating families about the visit to continually gather stakeholder perspectives. In situations when data collection was less successful than expected, caregivers and staff reflected together about challenges and potential future approaches. Families were offered a second visit in the event that insufficient data were obtained at the first visit.

## Results

To assess acquisition success, we used separate metrics for EEG and ET as participants varied in their abilities to wear the EEG net and complete the eye-tracking calibration procedure. EEG preprocessing included (1) data filtering from 1-45 Hz; (2) bad channel identification and interpolation; (3) wavelet denoising; (4) segmentation into 1s epochs; (5) artifact detection. Segments with greater than 10 channels exceeding 120uV were considered artifactual. Similar criteria have been used in prior literature including children with ID (e.g., DiStefano et al., 2019; Proteau-Lemieux et al., 2021; Saby et al., 2022). Additionally, any recording with more than 64 bad channels prior to segmentation was considered EEG acquisition failure. Data acquisition success for EEG was measured by whether the participant contributed 7 minutes (840 epochs) of artifact-free trials. Data acquisition success for ET was measured by whether the participant attended to the screen for at least 65% of stimulus presentation time averaged across the static-scene presentations. ET processing included: (1) Segmentation of ET data into epochs subtending distinct static scene trials; (2) measuring the percentage of ET samples where attention was directed to the screen; (3) measuring the percentage of on-screen samples for each region of interest (e.g. faces) for each trial.

Our approach supported effective acquisition of EEG and ET data. 76 participants with ASD+ID (*M_age_*=8.76 years, *SD*=4.05; *M_IQ_*=32.4, *SD*=15.5) and 8 participants with ID only (*M_age_*=8.91 years, *SD*=3.72; *M_IQ_*= 45.9, *SD*=15.6) have participated to date. For EEG, net application was successful in 67% of the children with ASD+ID and 87% of the children with ID only. Usable data, defined as more than 7 minutes of segments was collected in 61% of the children with ASD and 88% of the children with ID. For ET, sufficient ET data, defined as more than 65% of onscreen looking time, averaged across static scenes presentations was achieved in 67% of children with ASD+ID and 63% of children with ID. This data acquisition rate is comparable to those for other studies from our group, including the ABC-CT multi-site study (Tsang et al., 2023), for autistic participants with IQ ranging from 60 to 130.

## Discussion

With this protocol, we demonstrated the viability of a novel EEG-ET data collection infrastructure and individualized behavioral supports not only for successful data collection but for quality research participation experiences for those with ASD+ID and profound autism. The approach described here leveraged existing behavior support protocols developed for individuals without ID (ABC-CT; McPartland et al., 2020), expertise of clinical staff, lived experiences of autistic people and their caregivers, and experiential knowledge research staff gained from participant visits. Each successive visit offered an opportunity to incorporate family feedback and iteratively refine procedures.

The innovation of an adaptive EEG-ET paradigm with customizable stimulus delivery allowed for at least partial data collection with the majority of participants, many of whom were participating in research for the first time. Flexible stimulus delivery also promoted data quality and maximized data collection time. Individualization and flexibility are important when conducting research with any clinical population, but especially for individuals with ASD+ID given limitations in functional communication, variable attention patterns, and high behavioral support needs. We utilized these guiding principles to prepare, anticipate, and remain as flexible as possible within the parameters for data collection, individualized for each participant. Research staff were prepared for unpredictability despite best laid plans and adapted and altered behavioral support and data collection approaches on a moment-by-moment basis. The order of visit events could be re-arranged in the event a child needed more time to warm up to the lab environment and build rapport with staff. Research staff aligned with participant needs and desires to implement an effective visit plan. Participants were provided as many choices as the experimental protocol allowed and as caregivers advised to maximize cooperation. Choices were presented in visual supports, seating arrangements, net desensitization, preferred activities, and behavioral reinforcers. Staff also anticipated the possibility of reinforcer satiation and had novel backup options.

We used applied behavior analysis-informed behavioral strategies in our research protocol but did not implement them with the intent of clinical intervention, which was beyond the scope of this research study. In line with standard best practice and ethical guidelines, principles of reinforcement were used to promote desired behaviors and to make the experience as enjoyable as possible for participants. No punishment procedures were utilized at any point during the study. Participants were free to engage in any form of behavior, but reinforcement was provided for desired behaviors. While watching preferred videos on screen was gaze-contingent, other behavioral supports were delivered non-contingently to minimize boredom and frustration, including distraction, sensory stimulation, and fidgets.

Beyond facilitating data collection, this work highlights the importance of designing research workflows to be accommodating of individuals with high support needs, balancing feasibility and flexibility. The interactive paradigm and accompanying behavioral supports were designed to make participation engaging and efficient, thereby increasing participation potential for individuals with higher autism severity and greater intellectual and communicative impairment; however, sessions were occasionally discontinued early due to distress and/or safety concerns.

Additionally, the interactive paradigm was devised to promote data quality through improved signal to noise ratio, but participant dysregulation could have impacted signal. Using a behavioral protocol for EEG data collection in a similar sample, DiStefano and colleagues (2019) found that participant state (e.g., level of agitation) at the time of EEG recording was associated with data retention regardless of age and intellectual ability. One potential solution to be explored in future work is the addition of autonomic arousal measurements through wearable devices during EEG sessions. Researchers have attempted to study autonomic arousal with heart rate variability and galvanic skin resistance in similar populations to some success and this is also being incorporated into clinical trials with non-speaking autistic patients (Kildal et al., 2023) and in combination with EEG (Saffaryazdi et al., 2022). As such, integration of real-time autonomic arousal measurement may provide valuable information about participant stress in future studies, including autistic participants who have ID/are minimally-to-nonspeaking.

Findings suggest that many research participation barriers can be reduced through proactive planning, individualized accommodations, and flexible implementation. Although research participation does not inherently provide direct clinical benefit, the approaches described here may also inform efforts to improve accessibility and tolerability in related settings, including healthcare, educational, and community environments. More broadly, this protocol, as well as those described by Tager-Flusberg et al. (2017) and Guy et al. (2021), may serve as practical frameworks for increasing inclusion of autistic individuals with intellectual disability in both research and clinical practice.

### Limitations and future directions

Importantly, our described methodology is limited in a few ways. Supporting study participation for individuals with intellectual and developmental disabilities can be more staff-, time-, and resource-intensive. Successful implementation of this protocol requires approximately 2-3 trained staff members in the EEG space. Training research staff to competency can take several months, particularly for those without prior experience working with autistic individuals. Additionally, access to specialized behavioral expertise, such as BCBAs and staff psychologists, may be limited in some research settings. Moreover, the participant-centered approach of this protocol relies on extensive, individualized visit preparation, including caregiver interviews, environmental preparation, and participant-specific behavioral reinforcer planning.

This data collection protocol was designed for a single session, thus requiring net desensitization to unfold rapidly. Additional acclimation visits to the lab or beginning procedures with caregivers or current therapists would allow for more gradual desensitization; however, attempts to desensitize outside of the lab and in the absence of trained staff may inadvertently increase an individual’s distress or aversion toward aspects of net wearing if not adequately responsive to an individual’s pace. Future work may consider systematically exploring individual factors associated with successful net desensitization in single session versus prolonged training approaches, as well as in laboratory versus external settings.

Participants in this study had idiopathic autism and thus were excluded for diagnoses of known genetic syndromes (e.g., fragile X syndrome, down syndrome). Given the high rate of ASD+ID/profound autism in these genetic syndrome populations, there is a need to expand our inclusive research protocol to these groups. New iterations of this study are in development to include individuals according to specific genetic syndromes.

### Conclusion

This framework for inclusive biomarker data acquisition builds on principles of community participatory research and aims to remain both transparent and open to continued participant and caregiver feedback. The direct inclusion of individuals with ASD+ID in quantitative research is vital not only for advancing knowledge of etiology and interventions, but also for refining research supports to enable more of this work to be conducted across institutions.

## Data Availability

All data produced in the present study are available upon reasonable request to the authors

