## Appendices A-E for "An Adaptive Technical and Behavioral Approach for Biomarker Acquisition in Autism with Intellectual Disability"

Appendix A. Parent interview

The goal of this call is for us to talk about what to expect during your visit and for me to ask some questions about CHILD, such as his/her likes or dislikes, and how he/she might respond to different parts of the visit so that we can create a fun and supportive environment for CHILD during his/her visit.

*Any additional questions to facilitate conversation, support rapport, or add additional/follow-up questions when appropriate.

1. **In appropriate detail describe the visit schedule and what to expect. Determine with parent what order or activities might be most comfortable and successful.**
    a. Sign consents and relax in private waiting room
    b. EEG Session (e.g., watch videos, wet net on head, parent in nearby waiting room unless needed in EEG room, etc.)
    c. Break in private lunch room
    d. Clinical assessment with clinician
    e. Behavioral observation with parent and/or clinician
2. **Note any parent questions or notes following discussion of expectations**
3. **Participant characteristics:**
    a. How would you classify CHILD'S language level? How does he/she communicate what he/she wants or needs? Are there any particular communicative behavioral cues he/she might give that we should be attuned to?
    b. How does CHILD do with meeting new people? How long does it typically take him/her to warm up?
    c. How does CHILD respond to single step verbal instructions (e.g. watch the screen, sit still)?
    d. How does CHILD respond to different sensory experiences? How might CHILD respond to a wet EEG net on his/her head?
    e. How does CHILD do with hair washing? Haircuts?
    f. How does CHILD typically act when he/she must separate from parent/caregiver?
    g. How does his/her behavior change when parent/caregiver is not in the room? (Establish a sense of whether study team should start EEG with parent/caregiver present)
4. **Selecting videos:**
    a. Does CHILD enjoy watching videos?
        i. If answer is no, ask about music
    b. Are there any specific YouTube videos CHILD really enjoys?
    c. How does CHILD typically act while watching these videos (e.g., gets excited, dances, sits still and watches video)?
        i. If possible, try to find videos that will be reinforcing and grab attention without increasing movement
5. **Reinforcement, visual support, and break time activities:**
    a. When CHILD is doing work at home or school, does he/she earn specific reinforcers?
    b. What other activities does CHILD do when he/she has a break or down time?
    c. Does he/she use any specific reward systems or other supports (e.g. token board, break timer)?
        i. Are you able to bring these with you to the study visit?
    d. When CHILD is doing work at home or school does he/she use visual supports (e.g., social stories, visual schedules)?

        i. Note: try to determine what visual supports if any may be useful during the study visit

6. **Potentially challenging behaviors:**
    a. Are there situations that might be frustrating or be overly stressful for CHILD?
    b. When CHILD becomes stressed or frustrated, how does he/she typically act?
    c. Is CHILD toilet trained? How might we know when he/she has to use the restroom?
        i. If not, does CHILD wear pull-ups or diapers?
    d. Is CHILD ever aggressive toward others?
        i. If yes, inquire about specific behaviors, antecedents, frequency, etc.
    e. Does CHILD ever try to harm himself/herself?
        i. If yes, inquire about specific behaviors, antecedents, frequency, etc.
    f. Does CHILD ever run away (e.g., out of room, out of building)?
        i. If yes, inquire about specific behaviors, antecedents, frequency, etc.
    g. Are there any other behaviors that we should be aware of during the study visit?
    h. How would you recommend we respond to these behaviors?
    i. How does CHILD typically calm down when he/she is upset and exhibiting these behaviors?
    j. Is there a behavior plan that staff implements at school? Would any aspects of that plan be helpful for us to implement for the visit?
    k. Are there any considerations related to medication for specific behaviors that we should be aware of and work with when scheduling the visit (e.g., best focus first thing in the morning, medication beginning to "wear off" at a certain time)?

7. **Additional notes and questions:**
    a. Does CHILD hold a pencil and write or draw?
        i. Determination of appropriate battery of clinical measures and if coloring can be a helpful activity to offer during netting to keep hands busy
    b. Do you have any recent IEPs or evaluation reports that you can share with us ahead of time to help us learn more about CHILD and prepare for his/her visit?
    c. Any additional notes
8. Proceed to any other pre-visit screening documents (e.g., diagnostic history, medical history)

Appendix B. Visual Support Documents

**Social Stories**

Customized social stories were provided to families ahead of their research visit to build children's familiarity and comfort with each portion of their session, including the EEG netting process. Each social story featured images paired with short, one-phrase descriptions of each step in the experimental protocol (e.g., "Put on towel," "Hat on head"). Stories also included photos of the research staff the child met on the day of their visit, as well as photos or graphics of prizes, breaks, and a visual schedule summarizing the steps of the session. Images and graphics were tailored to the family's visit based on the specific personnel involved and the child's preferred break activity, snack, and prize. At the start of the research visit, research personnel re-read the social story with the child to reinforce familiarity, provide predictability, and ease the transition into study activities.

**Visual schedules, token boards, first-then boards, and checklists**

Staff leveraged preferred characters, activities, and items to create customized visual supports and provide a framework of what to expect throughout the research visit. The type and modality (e.g., paper, iPad, white board) of visual support were determined based on caregiver report of previously effective strategies.

If a child was motivated by reward systems, they could earn tokens or stars after completing each step of the experimental protocol. For example, a participant who utilized token boards and visual schedules at school and had a strong interest in a major city's public transit system received a laminated bus shaped token board with bus pass velcro tokens to be placed after completing each step of the protocol. A sample customizable first-then board template is available for download.

| | |
|---|---|
| 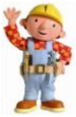 First… 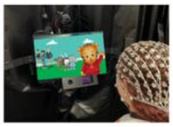 Put on hat and watch movies | Then… 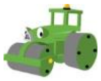 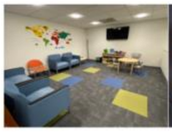 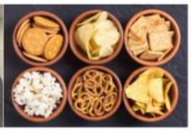 Break with snack |
| First… | Then… |

Appendix C. Parent/Caregiver Support Document (FULL)

Guide to Your Child's EEG Session

**What is EEG?**
Electroencephalogram (EEG) is a method to measure the electrical activity that our brains are naturally generating every second! While there are a few different types of EEG equipment, we use a net made of plastic nodes containing small sponges connected by rubber bands. During the process, we wet the sponges with a solution of water, salt, and baby shampoo to allow us to measure electrical activity in the brain. EEGs are very safe, but we understand that they may elicit some anxiety. We want to make the experience as positive for your child as possible! Here's how you can help:

**Before you arrive:**

We'll ask you a series of questions to help us anticipate how your child may react to the EEG and how we can make the experience as successful and positive as possible. These questions may include:

- How does your child do during haircuts, teeth brushing, and other similar activities?
- Do you think your child would be more comfortable with you in the room or in the waiting room?
- What kind of visual aids might your child benefit from, if any?
- What kinds of rewards or reinforcers might your child benefit from?
- How do you think your child would do with taking breaks? Would it be hard for them to return to the task?
- Are there any potential behaviors that you would like us to watch for? Indicators that your child is becoming upset?
- Are there any external events, items, etc. that may distress your child?

**When you arrive:**
We'll ask you a series of questions related to how your child's day is going and for any reinforcers or activities you brought with you

**Strategies to help your child feel as comfortable as possible may include:**
- Showing and letting your child touch the net
- Using a teddy bear to show your child how will put the net on their head
- Providing lots of praise and reinforcement when your child tolerates the net
- We can use a token board, stickers, snacks, or other reinforcers you think will help your child
- Re-reading the social story together slowly
- Looking over a visual schedule

- Reassuring your child that nothing will hurt

*Our goal is to go at your child's pace and not to force anything.*

**Behavior Management during the EEG**

Our experienced team, informed of your child's specific needs, will do all that we can to ensure a positive experience for your child. Based on what we decide together, you may begin either relaxing in the waiting room or come with us into the EEG space. If you begin in the waiting room, one of our staff will come and notify you should we need your assistance.

In the event that you are called in to assist with behavior management

- Try to focus on what you'd like your child to do (**"Hands down"**) instead of what you don't want them to do ("Don't touch the hat").
- Other helpful phrases to use only as needed include:
    - **"Sit quietly"**
    - **"Talk later"**
    - **"Hands in lap"**
    - **"Calm body"**
    - **"Watch screen"**
    - **"First movies, then prize"**
    - **"Great sitting still!"**
    - **"Awesome watching!"**

- Ideally, we hope to minimize the amount of talking during your child's session.
- Ideally, refrain from prompting your child to "look" or pointing to screen, but a simple and infrequent **"Watch screen"** is ok when the movies are paused.
- The behavioral support staff will guide the session and prompt your child as necessary.
- Provide positive reinforcement by praising your child for desired behavior (**"Nice job sitting still! Great work keeping the hat on!"**).

Appendix D. Parent/Caregiver Support Document (ABBREVIATED)

EEG Session Information for Caregivers

- Your child is likely to look to you to understand this unfamiliar situation. Modeling calmness and positivity helps many children to feel more comfortable.
- Focus on what you'd like your child to do (**"Hands down"**) instead of what you don't want them to do ("Don't touch the hat").
- Other helpful phrases to use only as needed include:
    - **"Sit quietly"**
    - **"Talk later"**
    - **"Hands in lap"**
    - **"Calm body"**
    - **"Watch screen"**
    - **"First movies, then prize"**
    - **"Great sitting still!"**
    - **"Awesome watching!"**
- Please minimize the amount of talking during EEG recording
- Ideally, direct your child's attention with a simple and infrequent **"Watch screen"** when the movies are paused. Refrain from prompting your child to "look" or pointing to screen.
- The behavioral support staff will guide the session and prompt your child as necessary.
- Provide positive reinforcement by praising your child for desired behavior (**"Nice job sitting still! Great work keeping the hat on!"**).

### Appendix E. Debrief Run Log

**Debrief Runlog**

*The purpose of this run-log is to provide both objective, quantifiable data (e.g., activity-level checkboxes) for analysis and descriptive session information for clinical feedback.*

| Participant and Visit Information | | | |
|---|---|---|---|
| **Study ID** | **Date** | **Time Point** | **EEG/ET Data Collected?** |
| ____________________ | ______________ | ◯ Day 1<br>◯ Day 2 | ☐ EEG Collected<br>☐ ET Collected<br>☐ Some EEG Collected<br>☐ Some ET Collected<br>☐ NO EEG Collected<br>☐ NO ET Collected |

1. **Child First Impression:**

◯ = One response only

☐ = Multiple responses acceptable

| **Context:** | **Observations:** | | |
|---|---|---|---|
| Waiting room | **Behavioral observations:**<br>☐ Interest in toys<br>☐ Interest in objects<br>☐ Interest in screens<br>☐ Interest in staff members<br>☐ Interest in parents<br>☐ Talking/saying words<br>☐ Vocalizing<br>☐ Crying<br>☐ Aggression towards others<br>☐ Self-injurious behaviors | **Child Affect:**<br>◯ Happy<br>◯ Neutral<br>◯ Irritable<br>◯ Sad | **Child activity level:**<br>◯ Low<br>◯ Medium<br>◯ High |
| Desensitization | **Behavioral observations:**<br>☐ Visual interest in net<br>☐ Proactively touching net<br>☐ Allowing net on head<br>☐ Pushing away net<br>☐ Crying/fussing when net is on/near head<br>☐ Aggression towards others when net is touched to head<br>☐ Self-injurious behaviors | **Child Affect:**<br>◯ Happy<br>◯ Neutral<br>◯ Irritable<br>◯ Sad | **Child activity level:**<br>◯ Low<br>◯ Medium<br>◯ High |
| Measuring head | **Behavioral observations:**<br>☐ Visual interest in tape<br>☐ Allowing tape on head<br>☐ Pushing away tape<br>☐ Crying/fussing when tape is on/near head | **Child Affect:**<br>◯ Happy<br>◯ Neutral<br>◯ Irritable<br>◯ Sad | **Child activity level:**<br>◯ Low<br>◯ Medium<br>◯ High |

| | |
|---|---|
| | ☐ Aggression towards others when tape is touched to head<br>☐ Self-injurious behaviors |

2. **Clinician and Parent Support during EEG/ET:**

| **Clinician/Parent Presence:** | **Response:** | | | |
|---|---|---|---|---|
| ○ Caregiver in the room<br>○ No caregiver in the room | **Time entered in room:**<br>○ Entered room with child<br>○ Entered room for netting<br>○ Entered during experiment | **Duration in room:**<br>○ Left after child sat<br>○ Left after netting<br>○ Stayed for entire experiment | **Parent Location:**<br>○ Sitting next to child<br>○ Child in parent's lap<br>○ Standing<br>○ N/A | **Caregiver:**<br>○ Mother<br>○ Father<br>○ Other |
| ○ Clinician in the room<br>○ No clinician in the room | **Time entered in room:**<br>○ Entered room with child<br>○ Entered room for netting<br>○ Entered during experiment | **Duration in room:**<br>○ Left after child sat<br>○ Left after netting<br>○ Stayed entire experiment | | |
| ○ Session called off<br>○ Session not called off | **Time session called off:** | **Person who made the call:**<br>○ Clinician<br>○ Caregiver | **Reason(s):** | |
| ○ Caregiver interference in study paradigm<br>○ NO Caregiver interference in study paradigm<br>○ N/A | **How much did it negatively impact the quality of the session?**<br>○ Low<br>○ Medium<br>○ High | **Invalidated trials?**<br>○ Yes<br>○ No | | |

3. **Equipment and Paradigm Setup:**

| **Event:** | **Response:** | |
|---|---|---|
| ○ EEG netting attempted<br>○ EEG netting not attempted | **Amount of attempts:**<br>○ 1<br>○ 2<br>○ 3<br>○ 4 | **Successful placement?**<br>○ Yes<br>○ No<br><br>**Net kept on for the whole session?** |

| | | | |
|---|---|---|---|
| | ◯ 5 | ◯ Yes<br>◯ No | |
| ◯ Impedances done<br>◯ No impedances done | **If not, why?** | | |
| ◯ ET calibration/validation attempted<br>◯ No ET calibration/validation attempted | **Amount of attempts:**<br>◯ 1<br>◯ 2<br>◯ 3<br>◯ 4 | | **Calibration/validation successful?**<br>◯ Yes<br>◯ No |

4. **Behavior during session:**

| **Behavior:** | **Response:** | |
|---|---|---|
| ◯ Not sitting<br>◯ Sitting | **How much did it negatively impact the quality of the session?**<br>◯ Low<br>◯ Medium<br>◯ High | **How effective was the response(s)?**<br>◯ Low<br>◯ Medium<br>◯ High |
| ◯ Net grabbing<br>◯ No net grabbing | **How much did it negatively impact the quality of the session?**<br>◯ Low<br>◯ Medium<br>◯ High | **How effective was the response(s)?**<br>◯ Low<br>◯ Medium<br>◯ High |
| ◯ Head moving<br>◯ Kept head still | **How much did it negatively impact the quality of the session?**<br>◯ Low<br>◯ Medium<br>◯ High | **How effective was the response(s)?**<br>◯ Low<br>◯ Medium<br>◯ High |
| ◯ Taking the sticker off<br>◯ Kept sticker on | **How much did it negatively impact the quality of the session?**<br>◯ Low<br>◯ Medium<br>◯ High | **How effective was the response(s)?**<br>◯ Low<br>◯ Medium<br>◯ High |
| ◯ Jerking body, arms, or legs<br>◯ No jerking | **How much did it negatively impact the quality of the session?**<br>◯ Low | **How effective was the response(s)?**<br>◯ Low<br>◯ Medium |

| | | |
|---|---|---|
| | ◯ Medium<br>◯ High | ◯ High |
| ◯ Kicking<br>◯ No kicking | **How much did it negatively impact the quality of the session?**<br>◯ Low<br>◯ Medium<br>◯ High | **How effective was the response(s)?**<br>◯ Low<br>◯ Medium<br>◯ High |
| ◯ Leaning/reaching (i.e., reaching for items off-screen, reaching for the eyetracker)<br>◯ No leaning/reaching (i.e., reaching for items off-screen, reaching for the eyetracker) | **How much did it negatively impact the quality of the session?**<br>◯ Low<br>◯ Medium<br>◯ High | **How effective was the response(s)?**<br>◯ Low<br>◯ Medium<br>◯ High |
| ◯ Stimming<br>◯ No stimming | **How much did it negatively impact the quality of the session?**<br>◯ Low<br>◯ Medium<br>◯ High | **How effective was the response(s)?**<br>◯ Low<br>◯ Medium<br>◯ High |
| ◯ Crying<br>◯ No crying | **How much did it negatively impact the quality of the session?**<br>◯ Low<br>◯ Medium<br>◯ High | **How effective was the response(s)?**<br>◯ Low<br>◯ Medium<br>◯ High |
| ◯ Not watching<br>◯ Watching | **How much did it negatively impact the quality of the session?**<br>◯ Low<br>◯ Medium<br>◯ High | **How effective was the response(s)?**<br>◯ Low<br>◯ Medium<br>◯ High |
| ◯ Self-injurious behaviors<br>◯ No self-injurious behaviors | **How much did it negatively impact the quality of the session?**<br>◯ Low<br>◯ Medium<br>◯ High | **How effective was the response(s)?**<br>◯ Low<br>◯ Medium<br>◯ High |
| ◯ Aggression<br>◯ No aggression | **How much did it negatively impact the quality of the session?**<br>◯ Low<br>◯ Medium<br>◯ High | **How effective was the response(s)?**<br>◯ Low<br>◯ Medium<br>◯ High |

<table>
<tr><td>○ Intentional disrupting behaviors (i.e., closing eyes at prompts, putting cape over face)<br>○ NO Intentional disrupting behaviors (i.e., closing eyes at prompts, putting cape over face)</td><td>How much did it negatively impact the quality of the session?<br>○ Low<br>○ Medium<br>○ High</td><td>How effective was the response(s)?<br>○ Low<br>○ Medium<br>○ High</td></tr>
<tr><td>○ Falling asleep<br>○ Awake for the duration of the experiment</td><td>How much did it negatively impact the quality of the session?<br>○ Low<br>○ Medium<br>○ High</td><td>How effective was the response(s)?<br>○ Low<br>○ Medium<br>○ High</td></tr>
<tr><td>○ Not interested in preferred videos<br>○ Interested in preferred videos</td><td colspan="2">How much did it negatively impact the quality of the session?<br>○ Low<br>○ Medium<br>○ High</td></tr>
<tr><td>○ Not interested in study paradigm<br>○ Interested in study paradigm</td><td colspan="2">How much did it negatively impact the quality of the session?<br>○ Low<br>○ Medium<br>○ High</td></tr>
<tr><td>○ No snacks<br>○ Not interested in snacks<br>○ Interested in snacks</td><td colspan="2">How much did it negatively impact the quality of the session?<br>○ Low<br>○ Medium<br>○ High</td></tr>
</table>

| **Clinician/Staff Notes:** |
|---|
